# Strengthening government’s response to COVID-19 in Indonesia: a modified Delphi study of medical and health academics

**DOI:** 10.1101/2020.11.09.20228270

**Authors:** Yodi Mahendradhata, Trisasi Lestari, Riyanti Djalante

**Affiliations:** Department of Health Policy and Management, Faculty of Medicine, Public Health and Nursing, Universitas Gadjah Mada, Yogyakarta, Indonesia; Center for Tropical Medicine, Faculty of Medicine, Public Health and Nursing, Universitas Gadjah Mada, Yogyakarta, Indonesia; United Nations University- Institute for the Advanced Study of Sustainability, Tokyo, Japan

## Abstract

The Indonesian government has issued various policies to control COVID-19. However, COVID-19 new cases continued to increase and there remains uncertainties as to the future trajectory. We aimed to investigate how do medical and health academics view the Indonesian government’s handling of the COVID-19 and which area of health systems that need to be prioritized to improve government’s response to COVID-19. We conducted a modified Delphi study adapting the COVID-19 assessment score card (COVID-SCORE) as the measurement criteria. We invited medical and health academics from ten universities across Indonesia to take part in the Delphi study. In the first round, participants were presented with 20 statements of COVID-SCORE and asked to rate their agreement with each statement using five-point Likert scale. All participants who have completed the first cycle were invited to participate in the second cycle in which they had the opportunity to revise their answer based on results of previous cycle and to rank a priority of actions to improve government response. We achieved consensus for 5 statements, majority agreements for 13 statements and no consensus for 2 statements. The prioritization suggested that top priorities for improving government’s response to COVID-19 in Indonesia, according to medical and health academics, encompass: (1) The authorities communicate clearly and consistently about COVID-19 and provide public health grounds for their decisions; (2) Everyone can get a free, reliable COVID-19 test quickly and receive the results promptly; (3) Contact tracing is implemented for positive cases; (4) Public health experts, government officials, and academic researchers agree on COVID-19 nomenclature and clearly explain the reasons for public health measures; and (5) Government communications target the entire diverse population. Ultimately, our study highlights the importance of strengthening health system functions during the pandemic and to improve health system resilience for dealing with future public health emergencies.

## Introduction

The COVID-19 pandemic is an unprecedented global crisis in the 75-year history of the United Nations [1]. As of 25 October, over 42 million COVID-19 cases and 1.1 million deaths have been reported globally [2]. Most governments had not anticipated the rapid spread of COVID-19 spread and were mostly reactive in their crisis response [3]. Governments’ failures to suppress the epidemic is disappointing and costly. One reason for failure to prevent the spread of SARS-CoV-2 is medical populism; politicians downplaying the pandemic, promoting simplistic solutions, or popularizing their responses to crisis [4]. Many governments are over relying on the rapid development of a vaccine against COVID-19, while immediate priorities should go to well-tested public-health measures for controlling outbreaks [5]. Control measures must cover all health system building blocks, such as service delivery, medical products and technologies, health information systems, health workforce, financing leadership and governance [6]. COVID-19 response has clearly highlighted the need for governments to improve their outbreak response by incorporating a comprehensive health-systems approach.

Indonesia is the world’s largest archipelago and world’s fourth most populated country, with more than 260 million people; its size and diversity puts it in a challenging position that very few other countries face [7]. The first two COVID-19 cases in Indonesia were confirmed on March 1, 2020, two months after SARS-CoV-2 was first reported on December 31, 2020 in Wuhan, China [8]. The initial zero case reported by Indonesia prior to the first two confirmed cases, was questioned by many. During this period, the government did not issue any form of travel restrictions and specific quarantines of travellers coming in/coming back to Indonesia, despite reports of increasing numbers of COVID-19 cases in neighbouring countries [9]. The Indonesian government only issued a travel restriction on January 27, 2020 from Hubei province, while at the same time evacuated 238 Indonesians from Wuhan.

After the initial and subsequent reports of infections, the Indonesian government started to realize the seriousness of the situation [9]. Still, the regulation on the large scale social distancing that restrict non-essential population mobility was only enacted a month after the first cases were reported, and subsequently implemented in the capital, Jakarta, on April 10, 2020 [10]. The government has since issued various policies and actions to tackle COVID-19 such as appointing general hospitals as COVID-19 Referral Hospitals [9]. However, the trend of COVID-19 new cases continued to increase. By 25 October 2020, Indonesia has reported 392,934 cases and 774 deaths [2]. There remains uncertainties and concern as to the future trajectory of COVID-19 in the country [7]. The impact on Indonesia’s vulnerable health system will be devastating, if COVID-19 continues the trajectory as has been observed in other countries. In the meanwhile, the economic, social, and non-COVID-19–related health system impact has already taken a great toll on Indonesia.

Clearly, coordinated and comprehensive actions to supress the COVID-19 epidemic in Indonesia need to be enhanced. The government need to prioritise advice from medical and health professionals [4]. The overall aim of this study is to consolidate advises from medical and health professionals for the government to enhance COVID-19 response in Indonesia. The study objectives are twofold. First, to investigate how do medical and health academics view the government of Indonesia’s handling of the COVID-19 epidemic. Second, to identify which area of health systems that need to be prioritized to improve government’s response to COVID-19 and strengthen health system resilience. This manuscript describes the modified Delphi method employed to achieve the study objectives, presents consolidated assessments by medical and health professionals on government’s COVID-19 response, ranked priorities for improvement, and elaborates their implications.

## Materials and Methods

### Research design

We conducted a modified Delphi study to address the study objectives. The Delphi method is commonly use in developing consensual guidance on best practice and exploration of a field beyond existing knowledge and the current conceptual framework [11]. In the context of COVID-19 in Indonesia, there are ongoing debates whether the government is actually capable to control the epidemic, whether appropriate measures have been taken, is the health system responding in coordinated manner. Therefore, Delphi method is suitable to build consensus of experts on those issues.

We adapted the COVID-19 assessment score card (COVID-SCORE) as the measurement criteria for the modified Delphi study [5]. COVID-19 SCORE is a recently developed measurement tool that could be used to assess government accountability in managing Covid-19 epidemic or other disease outbreaks. The tool was created based on the elements of Pandemic Health System Framework, which was adapted from the WHO’s health system framework [6]. The COVID-SCORE consists of twenty policy statements about improving public health communication and health literacy, facilitating robust surveillance and reporting, developing pandemic preparedness, strengthening health system, ensuring health and social equity, and ensuring comprehensive confinement and de-confinement strategies. The twenty statements from COVID-SCORE were then modified into three types of questionnaires: Likert scale survey, short comment, and ranking scale. The investigators in this study reviewed the adopted statements, assessed content validity, construct validity and approved changes.

The adopted COVID-SCORE questionnaire was tested to six medical and health academics who are not working in the institutions participating in the study. Pilot test participants filled out the questionnaires and provided comments on face validity and clarity of statements in the end of pilot study. The questionnaire was then revised accordingly.

### Study population

We adopted Cambridge Dictionary’s definition of academic: a person who teaches in a college or university [12]. To be considered as participant in this study each candidate has to fulfil the following criteria: (1) Hold a master or PhD degree in public health or clinical fields, (2) A researcher or lecturer in a reputable public university, and (3) Working at least two years at the selected institution.

We aimed to have regional representativeness of academics in the panel. Thus, we started with purposively selecting potential universities to represent four regions of Indonesia: Sumatera, Java, Kalimantan, and Eastern Indonesia. More than half of Indonesian population reside in Java island and majority of universities are also located in Java. Thus, higher proportion of participants was allocated to universities in Java island. To ensure that respondents have the capacity and capability to participate in the survey, we asked senior lecturers to recommend a list of academics from their university.

The Delphi group size depend on group dynamics for arriving at consensus among experts rather than on statistical power [13]. Since Delphi method is an iterative online process, there is a high risk of non-respond participant at every phase of study. To ensure enough number of participants is available at the last stage, we recruited at least double the number of participants needed at the last stage (about 30). An overview of the recruitment process is presented in **Fig 1**.

**Fig 1.**
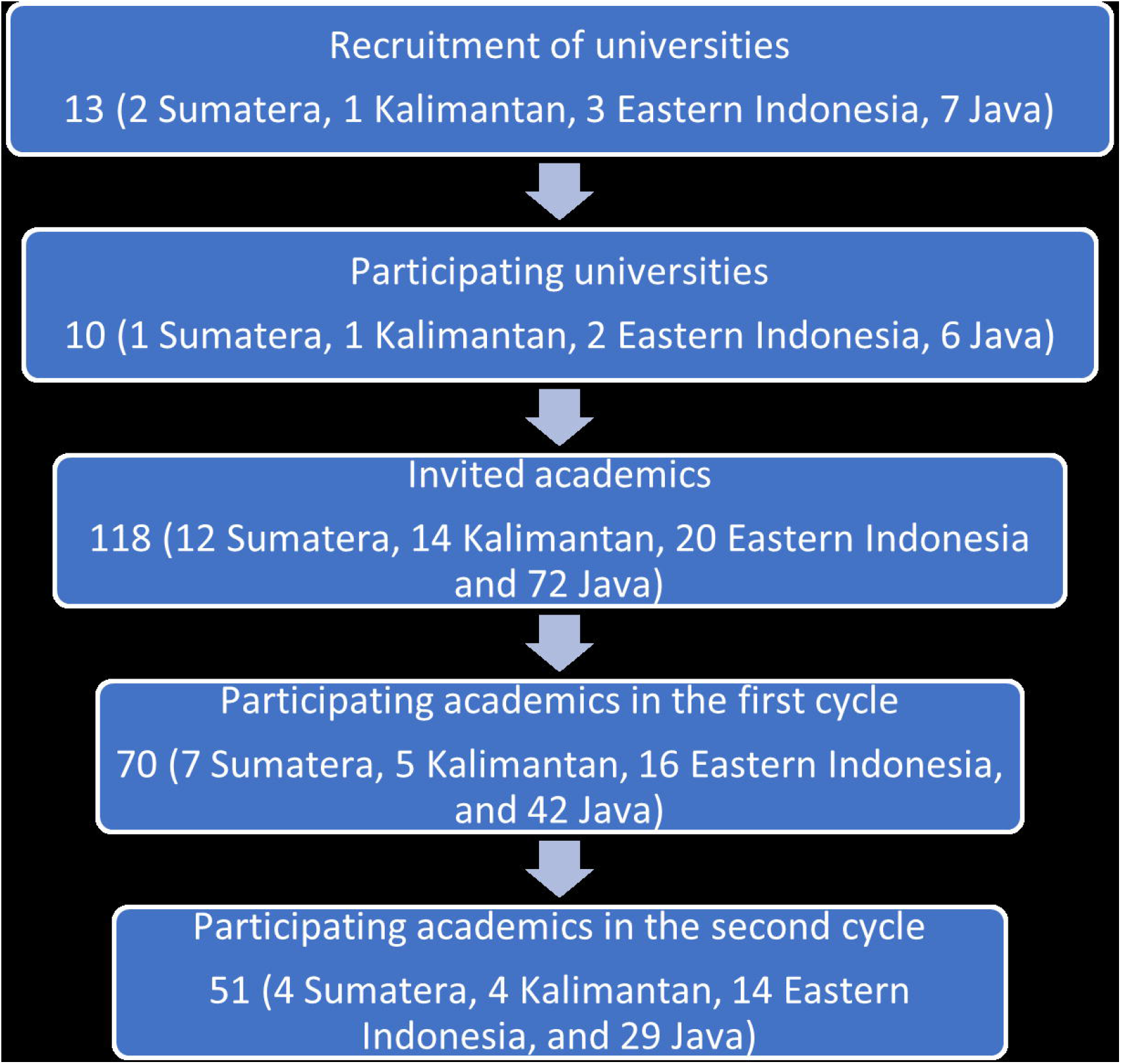
Recruitment process for the modified Delphi study

### Data collection

We sent invitation by email to 118 academics from 10 universities in Indonesia to participate in the first round of online Delphi Survey. The email contained a short description of the background and aims of the study, the process of two cycle Delphi survey, and a link to an online questionnaire on Qualtrics Research Suite platform.

In the first round, participants were presented with 20 statements of COVID-SCORE and asked to rate their agreement with each statement using five-point Likert scale: (1) completely disagree, (2) disagree, (3) neutral, (4) agree, and (5) completely agree. All statements were written in Bahasa Indonesia and in English. Participants could optionally add short comments or reflection relating to each statement in free text format. Participants had 7 days (20 to 26 July 2020) to fill out the questionnaire. A reminder was sent two days before the due date and we extended the first cycle for one day. A total 71 academics participated in the first cycle.

All participants who have completed the first cycle were invited to participate in the second cycle of Delphi survey which took place between 28 July to 3 August 2020. Summary of results from the first cycle was re-administered to all participants. Analysed data reflected more objective view on each statement and provided more confident and solid direction for panel review. After reading the results, participants had a chance to revise their original responses so the end results should be closer to a satisfactory degree of consensus. All experts in the panel were kept anonymous to each other, but not to the researcher, throughout the process. Based on the result of the first cycle, participants were asked to rank priority of actions to improve government response to COVID-19 pandemic. A reminder was sent two days before the due date and a final reminder was sent on the last day of data collection. Result of the second cycle was analysed and feedback to all participants a week later.

### Analysis

Participant profile, i.e. age, gender, profession, and region were coded. Percentages of score for each statement were calculated. “Strong Consensus” was defined as ≥95% agreement in the rating of the single statements by the panellists; “consensus” was defined as agreement of 75–94%, “majority agreement” was defined as agreement of 50-74%, and “no consensus” was recorded if the agreement was <50%. Agreement was if the panellists replied either “agree” or “strongly agree” and disagreement if “disagree” or “strongly disagree”. In the second cycle, respondent had the opportunity to revise their answer. Although it was not compulsory, the panellists also had the opportunity to provide comments on the statements or reasons for their assessment. For ranking score, each priority action will be scored according to its sum of rank. The lowest total score will be the highest priority. After completion of preliminary analysis, all experts in the panel were invited to a webinar to discuss the research brief which summarized the process and interpretation of findings from the Delphi survey. The research brief has also been shared with the National Planning Development Agency.

### Research Ethic

Informed consent was required before a participant could start filling the online questionnaire. Anonymity is one of the main features that characterized this study method. Participant did not meet with each other and did not get any information about other participants involved in the study. Therefore, they can freely submit their ideas, unbiased by identities or pressures of other. All participants were coded in the analysis; thus, their opinion and comments are also anonymous to the investigators. The study was reviewed and granted ethical approval by the Medical and Health Research Ethics Committee Faculty of Medicine, Public Health and Nursing Universitas Gadjah Mada/Dr. Sardjito General Hospital (Protocol ID KE/FK/0743/EC/2020).

## Results

Out of 118 academics invited to participate in this study, 75 responded to the invitation. Two individuals declined to participate and three did not finish the survey. In total, 70 academics from 10 public universities completed the first cycle of online Delphi survey (response rate 59%), ensuring representation from four Indonesia’s regions (Sumatera: 1 university, 7 participants (10.0%); Kalimantan: 1 university, 5 participants (7.1%); Eastern Indonesia: 2 Universities, 16 participants (22.8%); and Java: 6 universities, 42 participants (60.0%). Gender was equally represented in this study (35 males and 35 females). Age ranged from 27 to 71 years, and median 46 years. One third of participants hold a master degree (22, 31.5%) and the rest hold a doctoral degree (48, 68.5%). Most participants were from the public health field (48, 69.0%), and the rest were from clinical field (12, 16.9%), health nutrition (5, 7.0%), nursing (3, 4.2%) and biomedical science (3, 4.2%). Most participants (53, 75.7%) were not directly involved with COVID-19 response team. It was not compulsory for respondent to provide comments for each of the Likert statements, but 50 (71%) were willing to share their opinions.

We sent invitation to participate in the second cycle of Delphi survey to 68 respondents. Two respondents from the first cycle were excluded because they did not provide contact details. It was recorded that 58 respondents access the survey, but only 51 completed the survey (75.0% response rate). About 60% of respondent (31 individuals) revised their response to Likert scale questions and 47 (92%) proposed priority rank of actions.

Result of the online Delphi survey is summarized in **Table 1**. Out of 20 survey statements, we achieved consensus for 5 statements, 13 majority agreements and no consensus for 2 statements. Compare to the first circle, agreement was improved for 15 statements and reduced for 4 statements. Majority of respondent agreed to 11 statements and disagreed to 9 statements.

**Table 1.** Consensus on policy statements and ranked priorities

| No | COVID-SCORE statements | First cycle (N: 70) |  |  |  | Second cycle (N: 70) |  |  |  |  | p-value | Comments |  |  |  | Rank |  |
| --- | --- | --- | --- | --- | --- | --- | --- | --- | --- | --- | --- | --- | --- | --- | --- | --- | --- |
|  |  | Agreement | Neither agree nor disagree | Disagreement | Agreement / disagreement proportion | Agreement | Neither agree nor disagree | Disagreement | Agreement / disagreement proportion | Interpretation | Significance change | Total | Positive sentiment | Negative sentiment | Mixed sentiment | Total score | Rank |
| 1 | The authorities communicate clearly and consistently about COVID-19 and provide public health grounds for their decisions. | 26 | 10 | 34 | 48.5% disagree | 24 | 10 | 36 | 51.4% disagree | Majority agreement | 0.37 | 42 | 3 | 33 | 6 | 224 | 1 |
| 2 | Government communications target the entire population in all its diversity (e.g. language, culture, education, and socioeconomic level). | 20 | 10 | 40 | 57.1% disagree | 14 | 8 | 48 | 68.5% disagree | Majority agreement | 0.08 | 44 | 5 | 31 | 8 | 305 | 5 |
| 3 | Public health experts, government officials, and academic researchers agree on COVID-19 nomenclature and clearly explain the reasons for public health measures | 23 | 18 | 29 | 41.4% disagree | 19 | 13 | 38 | 54.2% disagree | Majority agreement | 0.07 | 37 | 6 | 20 | 11 | 288 | 4 |

|  |  | First cycle (N: 70) |  |  |  | Second cycle (N: 70) |  |  |  |  | p-value | Comments |  |  |  | Rank |  |
| --- | --- | --- | --- | --- | --- | --- | --- | --- | --- | --- | --- | --- | --- | --- | --- | --- | --- |
|  |  | Agreement | Neither agree nor disagree | Disagreement | Agreement / disagreement proportion | Agreement | Neither agree nor disagree | Disagreement | Agreement / disagreement proportion | Interpretation | Significance change | Total | Positive sentiment | Negative sentiment | Mixed sentiment | Total score | Rank |
| 4 | Everyone can get a free, reliable COVID-19 test quickly and receive the results promptly | 6 | 10 | 54 | 77.1% disagree | 7 | 8 | 55 | 78.6% disagree | Consensus | 0.41 | 42 | 1 | 39 | 2 | 267 | 2 |
| 5 | Contact tracing is implemented for positive cases | 46 | 11 | 13 | 65.7% agree | 53 | 9 | 8 | 75.7% agree | Consensus | 0.10 | 37 | 7 | 24 | 6 | 279 | 3 |
| 6 | Public health bodies maintain robust national, subnational, and local epidemiological databases, updated and reported daily | 47 | 10 | 13 | 67.1% agree | 50 | 11 | 9 | 71.4% agree | Majority agreement | 0.29 | 36 | 10 | 18 | 8 | 379 | 8 |
| 7 | There are enough qualified health workers and medical equipment (e.g. ventilators and face masks) to meet national needs | 21 | 10 | 39 | 55.7% disagree | 18 | 5 | 47 | 67.1% disagree | Majority agreement | 0.08 | 35 | 5 | 30 | 0 | 354 | 7 |
| 8 | The government can require private manufacturers to produce critical equipment rapidly, if needed | 50 | 10 | 10 | 71.4% agree | 50 | 9 | 11 | 71.4% agree | Majority agreement | 0.5 | 29 | 18 | 10 | 1 | 518 | 11 |
| 9 | A pandemic preparedness team that includes public health and medical experts is coordinating the national response | 40 | 16 | 14 | 57.1% agree | 39 | 14 | 17 | 55.7% agree | Majority agreement | 0.43 | 30 | 8 | 17 | 5 | 382 | 10 |
| 10 | Infection prevention and care guidelines and protocols are comprehensive and up to date | 41 | 21 | 8 | 58.6% agree | 43 | 20 | 7 | 61.4% agree | Majority agreement | 0.2 | 30 | 8 | 11 | 11 | 347 | 6 |
| 11 | Health systems have sufficient funding and infrastructure to care for all COVID-19 patients. | 13 | 17 | 40 | 57.1% disagree | 12 | 13 | 45 | 64.3% disagree | Majority agreement | 0.19 | 33 | 3 | 21 | 9 | 380 | 9 |
| 12 | Everyone has uninterrupted access to regular health services | 16 | 16 | 38 | 54.3% disagree | 14 | 11 | 45 | 64.3% disagree | Majority agreement | 0.11 | 34 | 4 | 27 | 3 | 554 | 13 |
| 13 | Primary care services and social services are coordinating and collaborating with each other during the pandemic | 25 | 12 | 23 | 35.7% agree | 23 | 23 | 24 | 34.3% disagree | No consensus | 0.43 | 30 | 10 | 16 | 4 | 562 | 14 |
| 14 | Mental health outreach services have been expanded to meet increased demand | 11 | 26 | 33 | 47.1% disagree | 11 | 12 | 37 | 77,1% disagree | Consensus | 0.00 | 28 | 8 | 19 | 1 | 760 | 20 |
| 15 | Task-sharing, task-shifting and telehealth are being used to optimize the delivery of health care services | 26 | 20 | 24 | 52% agree | 29 | 17 | 24 | 52.9% agree | Majority agreement | 0.46 | 29 | 6 | 17 | 6 | 651 | 18 |
| 16 | Appropriate measures have been taken to protect members of vulnerable groups such as the elderly, the poor, migrants, and the homeless | 16 | 16 | 38 | 54.3% disagree | 10 | 27 | 43 | 61.4% disagree | Majority agreement | 0.19 | 35 | 5 | 28 | 2 | 543 | 12 |
| 17 | COVID-19 efforts are focused on densely populated, low-resource areas | 23 | 20 | 27 | 38.6% disagree | 19 | 20 | 31 | 44.3% disagree | No consensus | 0.25 | 30 | 8 | 22 | 0 | 625 | 16 |
| 18 | Public health measures have been implemented to protect people in institutions and other confined settings | 34 | 15 | 21 | 61.8% agree | 33 | 10 | 27 | 55,0% agree | Majority agreement | 0.22 | 28 | 8 | 14 | 6 | 676 | 19 |
| 19 | The government is addressing the health and socioeconomic impacts of instituting and easing containment measures | 34 | 25 | 11 | 77.3% agree | 37 | 11 | 12 | 75,5% agree | Consensus | 0.41 | 28 | 7 | 16 | 5 | 611 | 15 |
| 20 | The government is collaborating with other countries, WHO, and other international bodies in responding to the pandemic | 48 | 13 | 9 | 84.2% agree | 49 | 14 | 7 | 87,5% agree | Consensus | 0.30 | 24 | 11 | 10 | 3 | 638 | 17 |

### Statements with consensus

Consensus was achieved for the following statements: (1) the government has maintained partnership with the WHO, other countries and international NGOs in responding to the pandemic (87.5% agree), (2)the government has tried to address the health and socioeconomic impact of instituting and easing containment measures (75.5% agree), (3) contact tracing is implemented for positive cases (75.7% agree); (4) everyone can get a free, reliable COVID-19 test quickly and receive the results promptly (78.6% disagree), (5) Mental health outreach services have been expanded to meet increased demand (77.1% disagree).

#### Government collaboration with other countries, WHO and international bodies

Most respondent agreed that the government has maintained partnership with the WHO, other countries and international NGOs in responding to the pandemic (consensus, 87.5% agree). However, several respondents criticized that the form of partnership is not clear and lack of publication about activities that has been done for the partnership. Range of collaboration was also not broad enough, and a respondent suggest to expand the collaboration with countries in Europe or United States. It was also suggested that adaptation of international guidelines or recommendations must be adjusted to local context and availability of resources in the country.

#### Contact tracing for positive cases

Majority of participant agreed that contact tracing has been implemented and it ranked third on priority action recommendation (consensus, 75.7% agree). We received 37 comments on contact tracing, out of it 28 respondents expressed their concerns regarding delayed initiation, coverage of contacts screened, unclear procedure, implementation variation at local level, coordination and resources needed when caseload continue to increase. There was also issue of patient honesty, and induced stigma due to staff visit to contact’s house. Six respondents mentioned that contact identification and tracings were not yet done for some positive cases.

#### Addressing the health and socioeconomic impacts of containment measures

Half of respondents agreed that the government has tried to address the health and socioeconomic impact of instituting and easing containment measures (consensus, 75.5% agree). The respondents were aware that the government has provided incentives and financial relief for vulnerable people who are affected by the pandemic. However, several respondents criticized that the measures were more focused on social and economic impact rather than health. Several respondents also suggested that weak monitoring and easing of confinement measures has created false perception that the situation is getting better. There was no sanction for people who did not wear face mask or did not adhere to infection prevention measures when in public.

#### Free, reliable COVID-19 test

Almost all respondents disagreed that COVID-19 test was available for free and result can be received promptly (consensus,78.6% disagree). Respondent acknowledge that COVID-19 test is available for free for those with COVID-19 symptoms or have immediate contact to COVID-19 patient. However, people who didn’t meet the diagnostic criteria must pay for any COVID-19 tests. At the beginning of the pandemic, diagnostic kits were scarce, and many people who wanted to be tested were rejected. Local government often conducted free mass screening but only in selected area, such as market and confined settings. When people get tested, often they had to wait for days or weeks before getting the test result. There were also concerns about sensitivity and specificity of rapid tests available in many healthcare facilities. Rapid test is often needed for administrative purposes, e.g. when people want to travel to another province, they need to show a rapid test result.

#### Mental health outreach services

Only 15% of respondents agreed that mental health outreach service was expanded during the pandemic (consensus, 77.1% disagree). Majority of respondent comments that they were not aware if such service is existing. However, many of respondent agreed that mental health outreach service is needed and should be expanded to reach people in high risk of suffering from mental illness due to the crisis. Information about availability of mental health service should be shared with the community.

### Statements with majority agreement

Majority of respondents agreed or disagreed that: the government has tried to maintain robust epidemiological databases at national and local levels (71.4% agree), the government should involve in country private manufacturers to produce critical equipment rapidly as part of corporate social responsibility (67.1% agree), a pandemic preparedness team that includes public health and medical experts is coordinating the national response (55.7% agree); infection prevention and care guidelines and protocols are comprehensive and up to date (61.4% agree); task sharing, task shifting and telehealth are being used to optimize the delivery of health care services (52.9% agree); public health measures have been implemented to protect people in institutions and other confined settings (55.0% agree), the government has target the entire population (68.5% disagree), there were enough qualified healthcare workers and medical equipment to meet national needs (67.1% disagree), the government has communicate clearly and consistently about COVID-19 (51.4% disagree), the government have enough funding and infrastructure to care for all COVID-19 patients, particularly in the long run (64.3% disagree), public health experts, government officials, and academic researchers agree on COVID-19 nomenclature and has clearly explain the reason for public health measures (54.2% disagree), access to regular health services was uninterrupted (64.3% disagree), and that appropriate measures have been taken to protect members of vulnerable groups, such as the elderly, the poor, migrants, and the homeless (61.4% disagree).

#### Updated and reported epidemiological databases

It was acknowledged that the government has tried to maintain robust epidemiological databases at national and local levels (majority agreement, 71.4% agree). It ranked 8 on priority recommended actions. There were 36 comments on this issue which mostly indicate that epidemiological data was available and accessible. Nevertheless, respondents also highlighted the importance of accuracy, validity, accessibility, transparency, consistency, timely reporting, synchronization, utilisation, presentation and interpretation of data. It is believed that these issues have created confusion in the community and affected public trust to published epidemiological data.

#### Private manufacturers roles in the pandemic

Respondents generally agreed that the government should involve domestic private manufacturers to produce critical equipment rapidly as part of corporate social responsibility (majority agreement, 67.1% agree). It ranked 11 on priority recommendation action. Respondents who provided comments also indicated that the central and local government has not optimized the opportunity to collaborate with private manufacturers. The initiative to involve private sector was considered to start too late. Concern on product quality was also raised, thus guidance and supporting policy are needed to attract investors to collaborate with the government and ensure public trust to products quality. A respondent suggest that focus should be put on manufacturing and expanding access to fast, efficient, accurate and affordable diagnostic tool for COVID-19.

#### Pandemic preparedness team

Majority of respondent agree that the pandemic preparedness team has included public health and medical experts (Majority agreement, 55.7% agree). It ranked 10 on the list of recommended action. Implementation, however, this is varied between regions, depending on local governments. Three respondents mentioned that involvement of public health experts only started a month before the survey, which was considered late. Respondents also identified public health experts who provided recommendation on personal basis and did not represent their institution. Recommendation from public health experts was not always accepted and resulted in policy that is inappropriate from public health point of view. Thus, to what extent does public health experts have been involved in the response team remains questionable. Several respondents highlighted the importance of improving coordination and collaboration of the response team with public health and medical experts, at national and local level.

#### Infection prevention and care guideline

Majority respondent agreed that infection prevention and care (IPC) guideline is available and updated (majority agreement, 61.4% agree). The Ministry of Health has released the 5^th^ revision of the IPC guideline when the survey was conducted. Comments on the IPC guideline was mainly related with frequent update of the guideline, but not followed with appropriate dissemination and training of the new guideline. Various institutions and professional organizations were also releasing IPC guideline, which potentially inconsistent with the national IPC guideline. Monitoring and supervision for the IPC implementation also had not existed, and in many areas, materials needed for infection control was scarce. All of these led to diverse application of IPC guideline in the field.

#### Task sharing, task shifting, and telehealth

Less than half of respondent agreed that task-sharing, task-shifting and telehealth were used in healthcare services (majority agreement, 52.9% agree). In many health facilities, particularly private health facilities, telehealth has been used for some time. However, respondents were concerned that policy to guide the implementation of a wide range of telehealth services in the country was not available. Reporting and recording of patient case using telehealth system was not standardized, and resources to run telehealth was not available in many areas, particularly in rural area. Payment for telehealth services was also another issue that need to be discussed by policy makers. While telehealth is generally understood, several respondents commented that task-sharing and task-shifting were not well implemented and rarely discussed.

#### Protection of people in institutions and other confined settings

Almost half of respondent agreed that public health measures have been implemented to protect people in institutions or other confined settings (majority agreement, 55.0% agree). Local government has published policy to guide people who work from office or other working places. Intervention includes mandatory face mask, social distancing, and hand washing. Several respondents mentioned that protection of people in institutions is depend on institutions policy, and implementation was varied. There was no evaluation whether the policy is implemented or not and there was no sanction for breaking the rules. One respondent highlights the importance of adjusting ventilation system to ensure safe air circulation in a building.

#### Communication of COVID-19

Half of respondent disagreed that the government has communicated clearly and consistently about COVID-19 (majority agreement, 51.4% disagree). Respondents acknowledged that the government has used various communication channel to provide information on COVID-19 to community. There are several official websites providing updated daily caseload data, policy and recommendations and become the main source of information. However, many respondents criticized inconsistencies of information, particularly in the early pandemic phase. There were ministries and institutions provided conflicting information and create confusion in the community. Information were often unclear, ambiguous and public health background of policy decisions was often not explained. All of these lead to various interpretation and confusion, even among highly educated people. Presentation of data was monotone and didn’t show a sense of urgency. Lack of coordination in the delivery of information was also felt. A respondent suggested that government need to create innovation in their way of delivering information, to ensure its rapid distribution to the whole population. Since the establishment of national response team, communication has been getting more coordinated, clearer and more consistent.

#### Communications target the entire population in all its diversity

Most respondent did not agree that the government has targeted the entire population (majority agreement, 68.5% disagree). Other than the fact that Indonesia is the largest archipelagic country in the world with hundreds of tribes and languages, respondents also raised the importance to deliver information to the poor and disable. Health campaign has been produced by various institutions, and several campaigns have been made using local language. Respondents suggested that language use for the campaigns should be understandable by people with low literacy and adapting local wisdom. Use of internet and social media to share information has discriminated population living in rural without access to internet or television. Some respondents also doubted that flow of information has reached the lowest level of community group (the neighbourhood or RT/RW). There was a concern that information only reached those with direct link to the response team. Thus, it is important to involve community or religious leader in delivering information.

#### COVID-19 nomenclature

More than half of respondent disagreed that public health experts, government officials, and academic researchers agreed on COVID-19 nomenclature and has clearly explained the reason for public health measures (majority agreement, 54.2% disagree). Respondents commented that many people did not understand the nomenclature, regarding the different terms, people without symptoms (OTG), people in surveillance (ODP) and patient on surveillance (PDP), and what level of surveillance entails for each category. This issue was not only faced by the community, but also by officials who worked directly for COVID-19. Another confusion started when the nomenclatures and definitions were updated into eight new terms in mid-July. There were also issue regarding terms use for infection control, disease transmission and treatment. The government need to create a strategy to effectively informed the community about the updated nomenclature and their use.

#### Availability of qualified health workers and medical equipment

Majority of respondent disagreed that there were enough qualified healthcare workers and medical equipment to meet national needs (majority agreement, 67.1% disagree). In every part of the country, medical personal protective equipment was still so scarce and price is skyrocketing after months of COVID-19. With increasing number of cases, hospital beds and ventilators will not be enough. The shortage of PPE and medical equipment is more pronounced outside Java, where more hospitals located in rural areas. Increasing number of infected health workers is concerning, especially when case load is also increasing. In previous outbreaks, there were special hospitals assigned to manage cases. Thus, many hospitals were not prepared to handle infectious disease pandemic. Many healthcare workers were not appropriately trained and competent to handle infectious disease patient.

#### Funding and infrastructure for COVID-19

Majority of respondent disagreed that the government have enough funding and infrastructure to care for all COVID-19 patients, particularly in the long run (majority agreement, 64.3% disagree). Several respondents commented that funding is available and that the government has tried to fulfil the need for health infrastructure, but the use of fund was hampered by bureaucracy. Delays in the payment of hazard incentives for frontline healthcare workers hints that the procedure to release the funds is still complicated.

#### Access to regular health services

Majority of respondent disagreed that access to regular health services was uninterrupted (majority agreement, 64.3% disagree). During the pandemic, many healthcare facilities limited their services, such as by reducing opening hours, reducing daily patient quota, postponing elective surgery, and lengthening the interval of follow up visit for chronic disease patient. On the other hand, many people were afraid to visit healthcare facilities for fear of exposure COVID-19 cases and stigma associated with COVID-19. Several respondents commented that access to healthcare was available as usual, but the community was not well informed about it.

#### Protection of vulnerable groups

Only ten respondents agreed that appropriate measures have been taken to protect members of vulnerable groups, such as the elderly, the poor, migrants, and the homeless (majority agreement, 61.4% disagree). The government have released financial incentives for the poor and encourage elderly people and young children to stay at home. Many non-governmental organizations and philanthropic raise funds to support those affected by the COVID-19 pandemic, including the poor, elderly, as well as frontline healthcare worker. But many respondents commented that specific intervention or campaign targeting vulnerable population is still inadequate and can be optimized.

### Statements with no consensus

There was no consensus on whether primary care services and social services are coordinating and collaborating with each other during the pandemic and whether COVID-19 efforts are focused on densely populated, low-resource areas.

#### COVID-19 efforts on densely populated, low resource area

Almost half of respondent disagreed that COVID-19 efforts are focused on densely populated, low resource area (no consensus, 44.3% disagree, 27.1% agree). Many respondents were not aware if the government has been focusing the efforts on this specific population. However, many agreed to this recommendation due to high risk of transmission in such setting. Implementation of this recommendation will depend on the local government decision.

#### Coordination and collaboration of primary care services and social services

Only one third of respondent agreed that there were coordination and collaboration between primary care services and social services (no consensus, 34.3 disagree, 32.8% agree). Many NGOs are actively involved in the COVID-19 response and primary healthcare facilities usually collaborate with social services in their coverage area. However, several respondent commented that the role of primary healthcare and social services can be optimized through better coordination and collaboration. The local government plays important role in building this collaboration, as shown in several provinces.

## Discussion

Our study highlighted consensus among Indonesian medical and health academics in regard to that: (1) the government has maintained partnership with the WHO, other countries and international NGOs in responding to the pandemic; (2) the government has tried to address the health and socioeconomic impact of instituting and easing containment measures; (3) contact tracing has been implemented for positive cases ; (4) Not everyone can get a free, reliable COVID-19 test quickly and receive the results promptly; and (5) Mental health outreach services have not been expanded to meet increased demand.

Our findings accordingly highlighted that the top priorities for improving government’s response to COVID-19 in Indonesia encompass: (1) The authorities communicate clearly and consistently about COVID-19 and provide public health grounds for their decisions; (2) Everyone can get a free, reliable COVID-19 test quickly and receive the results promptly; (3) Contact tracing is implemented for positive cases; (4) Public health experts, government officials, and academic researchers agree on COVID-19 nomenclature and clearly explain the reasons for public health measures; and (5) Government communications target the entire population in all its diversity (e.g. language, culture, education, and socioeconomic level). These recommendations are discussed in more details in the following paragraphs.

**Communication** is considered to be critical for effective government response by our Delphi study participants, highlighting three communications goals within the top five priorities (Priority rank 1;4 and; 5). There have been numerous government communication problems that have occurred across multiple countries, including Indonesia [14]. Ineffective risk communication for instance clearly impeded the emergency response in Wuhan’s outbreak management [15]. The Wuhan government did not integrate scientific risk communication into policy decision, the local government even delayed reporting and handled the information publicity in an ambiguous way which diminished public perception associated with COVID-19, and the government failed to manage uncertainty and different levels of risk perception of COVID-19. Communication discordance not only signifies the failure of governmental systems, which greatly diminishes public trust in the government but also considerably increases public confusion and fear about COVIDL19 risks [14]. It has often been challenging for the public to differentiate between evidence-based and less scientifically reliable information during the COVID-19 pandemic, partly due to poor communication by government officials [16]. Failure by governments to communicate effectively can seriously undermine their responses to COVID-19.

Arguably, Indonesia and many other countries have paid high costs due to COVIDL19, which could have been prevented and addressed much more effectively if governments had more responsive and strategic risk communication [14]. Communication is a substantial action, not just a precursor to an action [17]. Policy makers should cautiously consider the quality of information circulated through private sources and social networks [18]. Furthermore, when disseminating crucial health information, a variety of information sources should be used to ensure that heterogenous populations have timely access to essential knowledge. Governments thus need to improve their efforts on disseminating information on the pandemic, as well as employ strategies for improved communication management to citizens through social media as well as mainstream information sources [19]. As the epidemic evolves, merely sharing situation updates and policies may be inadequate to capture public attention [20]. Thus, governments should adopt a more inclusive and empathic communication style to address public concerns. Consistent, credible and targeted communication is critical in encouraging people to comply to COVID-19 control measures [16].

Our Delphi participants also emphasized the importance of **enhancing testing and contact tracing**. A test, trace and isolate strategy remains the most effective method of controlling the COVID-19 outbreak, until an effective vaccine has been developed [21]. The participants accordingly suggested that government should prioritize efforts to ensure that everyone can get a free, reliable COVID-19 test quickly and receive the results promptly. Timely and accurate laboratory testing is evidently essential to manage the COVID-19 pandemic [22]. Serological tests have the main shortcoming of a late positivity during the disease course, although attractive due to their lower cost and ease of implementation [22]. Many questions remain unanswered regarding the role of serological testing in COVID-19 diagnosis and monitoring. Reverse transcription-polymerase chain reaction (RT-PCR) thus remains the gold-standard for SARS-CoV-2 diagnosis, but several operational issues significantly limit the test use. Indonesia has been facing a hard time to rapidly scale-up laboratory capacity for RT-PCR [23]. Although the number of reference lab increases, the daily number of testing results remains fluctuated and unstable. This reflects operational challenges such as: readiness and capacity between labs; availability of swab collection officers; availability of reagents in the lab; rules for lab officers and swab collection officers; and transportation for a specimen from health facility to the referral lab.

In complement to enhancing testing, **contact tracing** for all positive cases should be a top priority for the Indonesian government, according to medical and health academics. Contact tracing prevent transmission of infectious diseases by identifying, assessing and managing people who have been in close contact with an infected individual [20]. A recent mathematical modelling exercise highlighted that contact tracing is a key component of the most rational scenario to control COVID-19 in Indonesia [24]. Another modelling exercise estimated that a high proportion of contacts need to be successfully traced to ensure an effective reproduction number lower than 1 in the absence of other measures [25]. Combined with moderate physical distancing measures, contact tracing and self-isolation would be more likely to achieve control of SARS-CoV-2 transmission. Contact tracing has been used extensively in previous emerging infectious disease outbreaks [26]. Recent studies suggested that the effectiveness of contact tracing could also be enhanced through app-based digital tracing [27]. The effectiveness of contact tracing and the extent of resources required to implement it successfully will ultimately depend on the social interactions within a population [28]. A recent review suggest that COVID-19 contact tracing systems could be facilitated by: clear communication about contact tracing; involvement of stakeholders in the development of contact tracing systems, particularly, digital applications; evaluation and quality assurance of the contact tracing system [21].

The key strength of this work was the ability to achieve consensus relating to government’s response to an ongoing pandemic from a considerable number of participants from multiple geographical regions across Indonesia. The process was done in the middle of a global pandemic over a relatively short period of time (six weeks), without the ability to hold face- to-face meetings.

Notwithstanding, our Delphi process had key limitations. Firstly, the scope of the work precluded inclusion of academics beyond medicine and health (e.g. economics, public policy, public administration) and non-academics (e.g. public health practitioners, clinical practitioners, policy makers, community leaders) in the process. Seeking opinions from a broader category of participants, would have been preferable had time and the need to ensure expertise of participants not been such pressing factors. Secondly, the statements assessed here were all developed based on expert opinions [5]. As the output and recommendations from a Delphi process can only be as robust as the statements that are assessed, higher levels of evidence and further experiences should be incorporated as they become available to ensure that the recommendations remains relevant.

## Conclusions

The COVID-19 pandemic has caused a global crisis that may endure for more than a year [29]. Citizens look to governments for leadership and credible information [30]. Pressure on governments to act decisively is enormous. To revamp COVID-19 response in Indonesia, the government urgently need to strengthen capacity to communicate clearly and consistently, based on commonly agreed COVID-19 nomenclature and target the entire diverse population. The government also need to ensure universal access to reliable COVID-19 test by expanding lab infrastructure and facilitating operational readiness. Finally, the government need to boost contact tracing implementation capacity and facilitate contact tracing for all positive cases, adopting tested innovative strategies (e.g. digital apps).

The recommendations from this Delphi study were primarily intended for the Indonesian government. Notwithstanding, governments of other countries may also benefit from this study by considering conducting similar exercise, utilizing modified Delphi method for rapidly assessing their response to COVID-19. Ultimately, our study highlights the importance of strengthening health system functions during the pandemic and to improve health system resilience for dealing with future public health emergencies.

## Supporting information

Supplemental reporting checklist

Supplemental codebook

## Data Availability

All relevant data are within the manuscript and its Supporting Information files.

## Acknowledgements

We would like to thank Ari Probandari, Bagoes Widjanarko, Chatarina Umbul Wahyuni, Dewi Susanna, I Wayan Gede Artawan Eka Putra, Mohammad Bakhriansyah, Rahayu Lubis, Sukri Palutturi, and Trevino Pakasi for their contribution to identify and recommend the potential participants for the modified Delphi study.

## Author Contributions

**Conceptualization:** Yodi Mahendradhata, Trisasi Lestari, Riyanti Djalante

**Formal analysis:** Trisasi Lestari

**Methodology:** Yodi Mahendradhara, Trisasi Lestari

**Investigation:** Yodi Mahendradhata, Trisasi lestari

**Supervision:** Riyanti Djalante

**Writing – Original Draft:** Yodi Mahendradhata, Trisasi Lestari

**Writing – review & editing**: Riyanti Djalante

**Supporting information**

**S1 Checklist. CREDES checklist**

(Docx)

**S1 Appendix. Delphi study data**

(Excel)

**S2 Appendix. Delphi study code book**

(Docx)

