## Supplemental reporting checklist for "Strengthening government’s response to COVID-19 in Indonesia: a modified Delphi study of medical and health academics"

CREDES Reporting Statement Checklist for

| *Checklist Item* | *Page no.* | *Description of how addressed in this manuscript* |
| --- | --- | --- |
| Purpose and rationale | 4-5 | The aim and objectives of the study have been explicitly presented in the end of the “introduction” section. The rationale for the choice of the Delphi technique has been presented in the first paragraph of the “method” section. |
| Expert panel | 6; 9 | Criteria for the selection of experts and information on recruitment of the expert panel have been reported in the “study population” sub-section of the “method” section. Characteristics of the study participants have been summarized in the first paragraph of the “result section.” Response rates over the ongoing iterations have been reported in the first and second paragraphs of the “result” section |
| Description of the methods | 5-8 | The methods have been described in detail, particularly in the “research design” and “data collection” sub-sections of the “method” section |
| Procedure | Figure 1 | The stages of the Delphi process have been summarized in Figure 1. |
| Definition and attainment of consensus | 8; 10 | Definition of consensus has been presented in the “analysis” subsection of the “method” section. The attainment of consensus has been reported in the “result” section |
| Results | Table 1; 15-25 | Details of the results have been summarized in Table 1 and key findings have been further elaborated in the “result” section. |
| Discussion of limitations | 28-29 | A critical reflection of potential limitations has been presented in the last paragraph of the discussion section |
| Adequacy of conclusions | 29 | The conclusions have been formulated based on the outcomes of the Delphi study and presented in the end of the manuscript |
| Publication and dissemination | 8 | Dissemination of the preliminary results have been described in the “analysis” sub-section of the “method” section |
