## Supplemental codebook for "Strengthening government’s response to COVID-19 in Indonesia: a modified Delphi study of medical and health academics"

COVIDSCORE

Nodes

| Name | Description |
| --- | --- |
| access to healthcare |  |
| access available |  |
| difficult to access |  |
| healthcare program affected |  |
| inconvenience for staff and patient |  |
| lack of information about access to HC |  |
| limited access to healthcare |  |
| People limit their visit to healthcare |  |
| service changes |  |
| varied between provinces |  |
| community response |  |
| COVID is thought as a common disease |  |
| Public trust to the government |  |
| People ignores safety measures |  |
| People won’t use personal protective devices |  |
| Community have different perspective and perception about the situation |  |
| contact tracing |  |
| coordination issue |  |
| delayed response |  |
| Difficulties in conducting contact tracing |  |
| limited resources for contact tracing |  |
| not optimal |  |
| Only symptomatic contacts are tested |  |
| not done |  |
| Not all contact were tested |  |
| patient honesty |  |
| Contact investigation was done |  |
| unclear procedures |  |
| not systematic |  |
| Coordination |  |
| Coordination between ministry, or between the central government with provincial and district government. |  |
| Different ways of coordination |  |
| Unclear roles and responsibility |  |
| Lack of public health expert involvement |  |
| Variation of public health involvement in the regions |  |
| Involvement of a public health expert as an individual and not on behalf on an institution |  |
| Need to improve partnership with health sector |  |
| Roles of public health is unclear |  |
| Public health expert is involved |  |
| Late involvement of public health expert |  |
| Public health expert was not involved in the decision making |  |
| Not all recommendation from the public health sector are heard and implemented. |  |
| Good collaboration between researcher, academician and the government is not shown. |  |
| Collaboration is varied at local level |  |
| Collaboration should be existed down to the neighbourhood level. |  |
| There is improvement in collaboration activity |  |
| There was no clear direction |  |
| Leadership was not seen |  |
| Not optimal |  |
| COVID Test |  |
| access varied |  |
| Unclear information |  |
| commercialization |  |
| delayed result |  |
| difficult access |  |
| free for suspect |  |
| Lack of capacity |  |
| larger scale |  |
| long queue |  |
| low sensitivity rapid test |  |
| not free |  |
| expensive |  |
| stigma for test takers |  |
| Database |  |
| Data accuracy |  |
| data error |  |
| synchronization |  |
| data is accessible |  |
| data is not feedback to grassroot |  |
| Data not transparent |  |
| Data utilization |  |
| utilization guideline |  |
| Delay reporting |  |
| inconsistent recording and reporting |  |
| lack of data interpretation |  |
| lack of detail |  |
| public trust |  |
| resources gap |  |
| focus on densely low resources area |  |
| Varied between regions |  |
| No policy |  |
| not optimal |  |
| lack of Human resources |  |
| Depend on the local government |  |
| Funding |  |
| Funding is available |  |
| Need funding for preventive measures |  |
| slow transfer of fund |  |
| Lack of transparancy |  |
| Limited use of fund |  |
| Varied use of fund in the provinces |  |
| inoptimal use of fund |  |
| Availability of fund in the long run is questionable |  |
| healthcare workers |  |
| Limited number of human resources |  |
| Need to improve incentive |  |
| incompetent volunteer or staff |  |
| lack of experience in managing pandemic |  |
| risk of infection among HCWs |  |
| uneven distribution of HCWs |  |
| infection control guideline |  |
| aplikasi di daerah bervariasi |  |
| Availability of guideline |  |
| Different guidelines made by various ministries |  |
| Guideline was adopted from the WHO |  |
| Need an SOP for communicating and disseminating the guideline |  |
| available |  |
| Many organizations are releasing covud-related guidelines |  |
| Too many updates and community can’t follow |  |
| The guideline can be improve |  |
| Many people don’t know about the IPC guideline |  |
| Need a monitoring and supervision plan |  |
| Lack of guideline dissemination |  |
| Difficult to read and understand |  |
| It does not support by availability of materials and other logistics at local level |  |
| Various available guidelines are not consistent |  |
| Some guidelines are not conformed to the WHO guideline |  |
| It is not clear if the guideline has been updated or not |  |
| International collaboration |  |
| We learn from policies from other countries |  |
| The collaborations are not clear |  |
| It must be done |  |
| Impact or result from the collaboration is |  |
| Results or impacts of collaborations are not clear |  |
| Collaboration more often is conducted with international NGOs |  |
| Collaboration is limited to certain countries |  |
| It’s not clear to what extend is the collaboration |  |
| There are collaborations with other countries |  |
| NGOs and private involvement |  |
| Not clear how its developed |  |
| Not optimal |  |
| There should be a quality guarantee |  |
| Should be collaborate to create an effective, effisien, fast and affordable devices. |  |
| Corporate Social Responsibility funding can be use |  |
| Need a specific policy |  |
| Private company also face economic challenges |  |
| Has been done to produce PPD |  |
| Depend on the local government |  |
| It started late |  |
| Mainstreaming local production |  |
| Collaboration between facility |  |
| Not optimal |  |
| Assisted by NGO |  |
| Lack of collaboration with private health facility |  |
| Reduce services in health facilities |  |
| Need a SOP for collaboration |  |
| There is a SOP for patient’s transfer between facilities |  |
| There is existing collaboration and coordination between facilities |  |
| Collaboration is better now |  |
| It is difficult to coordinate collaboration between facilities. |  |
| Communication |  |
| Communication style |  |
| Type of information is varied |  |
| False campaign |  |
| Weak public communication |  |
| Exclusive communication |  |
| Too simple |  |
| Negative findings |  |
| Lack of collaboration with community leaders and religious leaders |  |
| inequality to access to information |  |
| Only those who directly involved with COVID team have access to updated information |  |
| information doesn’t reach the community |  |
| Not everyone has access to social media/internet |  |
| Unclear roles |  |
| Information not equally distributed to community |  |
| Inefficient and ineffective information |  |
| There is not enough information related with evidences |  |
| Description is monotone |  |
| Unclear communication |  |
| Ambiguous sentence |  |
| Content doesn’t fit with local context |  |
| Lack of time for communication |  |
| Misunderstanding about COVID is still exist in the communicaty |  |
| People understanding is stuck with terminology. |  |
| Nomenclatures are not fully understood |  |
| Nomenclature are confusing |  |
| Nomenclature are often changing |  |
| Nomenclatures have been revised |  |
| Multiple interpretation |  |
| Inconsistent delivery of information |  |
| Can’t trigger public understanding |  |
| Inconsistent explanation |  |
| Between ministries, between central and local government |  |
| Government authorities released different statements |  |
| Difficult to generalize |  |
| Interventions were made based on local context |  |
| not evaluated |  |
| Public health recommendation is often not heard |  |
| Not reflects the urgency of the situation |  |
| Positive findings |  |
| Information spread fast |  |
| Information is available |  |
| Better information after formation of the national coordination team |  |
| Become the main reference |  |
| Use various media channel |  |
| Recommendations |  |
| Need technical assistance |  |
| Need enforcement |  |
| Need environment engineering |  |
| Need innovation in information channels |  |
| Source of information should be from one channel |  |
| tailored communication |  |
| Use local language or culturally sensitive language |  |
| No information for difable |  |
| Information should reach rural area |  |
| Consider various level of health literacy in the community |  |
| Use social media |  |
| no tailored communication |  |
| Communication topic was not comprehensif |  |
| Logistics |  |
| There are efforts to fulfill the logistics needs |  |
| distribution of facilities to healthcare |  |
| lack of HCF |  |
| lack of logistics |  |
| lack of PPE |  |
| not enough |  |
| overload risk |  |
| uneven distribution |  |
| Mental health |  |
| Online mental health service is available |  |
| Mental health service for covid-19 related cases is not available |  |
| Lack of mental health service for covid-19 |  |
| Different views about the interventions |  |
| policy |  |
| Dominated by few experts |  |
| Lack of priority for public health |  |
| Did not consider the situation in the community |  |
| Not evidence based |  |
| protection in confined settings |  |
| Not existed |  |
| Not evaluated |  |
| Not optimal |  |
| Varied implementation |  |
| Not strictly implemented |  |
| Workplace policy |  |
| Need to change the policy about room ventilation |  |
| Other than in health facility, it is non existing |  |
| Need a dissemination of the policy at workplaces |  |
| Several confined settings have become infection clusters |  |
| Many companies remains working from the office |  |
| stigma |  |
| Telemedicine, Task sharing and task shifting |  |
| Local application is varied |  |
| Only in certain professions |  |
| Not supported by policy |  |
| Staff do not understand about task sharing/shifting |  |
| Not available in rural area |  |
| not optimal |  |
| Type of services is very limited |  |
| Lack of technology |  |
| Less interaction with patient |  |
| Private health facilities can do it better |  |
| Expensive and not covered by health insurance |  |
| Still learning the software |  |
| No monitoring and evaluation mechanism |  |
| No reporting mechanism |  |
| Reduced infection |  |
| Community acceptance |  |
| There is task shifting |  |
| There is telehealth |  |
| Govt efforts to reduce the impact of restriction |  |
| Not seen yet |  |
| Limited financial relief |  |
| Missunderstanding of the program in the community |  |
| Need to involve stakeholders |  |
| Low social economy level should be prioritized |  |
| Weak monitoring and sanction |  |
| There is effort to provide relief |  |
| Inconsistent application |  |
| vulnerable groups |  |
| There is a financial relief |  |
| Transportation restriction for baby, child and pregnant mother |  |
| NGOs’ donation |  |
| Only as advice |  |
| Non existent |  |
| Not optimal |  |
| coverage and sustainability |  |
| Depend on the community |  |
| No support for homeless people |  |
| Threaten by loosening the restriction |  |
